# Assessing the Secondary Use and Scientific Impact of Shared Clinical Trial Data: A Cross-Sectional Study of Clinical Trials Shared on the YODA Project Platform

**DOI:** 10.64898/2026.03.26.26349328

**Authors:** Erfan Taherifard, Maryam Mooghali, Hollin R. Hakimian, Sahil R. Mane, Mengyuan Fu, Stephen Bamford, Jesse A. Berlin, Karla Childers, Nihar R. Desai, Cary P. Gross, Debbie Hewens, Richard Lehman, Jessica D. Ritchie, Tamsin Sargood, Joanne Waldstreicher, Joshua D. Wallach, Molly K. Willeford, Harlan M. Krumholz, Joseph S. Ross

## Abstract

**Objective:** To assess the number, timing of publication, characteristics, and scientific impact of secondary publications generated using individual participant-level data (IPD) from a portfolio of Johnson & Johnson-sponsored clinical trials shared with external investigators through a data sharing platform.

**Design:** Cross-sectional study.

**Setting:** Yale University Open Data Access (YODA) Project platform.

**Participants:** Johnson & Johnson-sponsored clinical trials listed on the YODA Project platform with IPD available for external sharing as of December 31, 2021, and with a full-length, peer-reviewed publication (i.e., primary publication) reporting primary endpoint results by the original trial investigators.

**Main outcome measures:** Number, timing of publication, research objectives, analysis type, and scientific impact of secondary publications using IPD from these trials identified through citation searches of primary publications in Web of Science through June 2025. Scientific impact metrics included journal impact factor, annual citation count, annual Altmetric Attention Score, and annual Mendeley reader count. Secondary publications were classified as internal (authored by at least one original trial investigator) or external.

**Results:** Among 336 eligible trials, 265 (78.9%) had at least one associated secondary publication, totaling 1,167 secondary publications, of which 209 (17.9%) were external. Among external secondary publications for which the data access mechanism was reported (n=190; 90.9%), most obtained access through data sharing platforms (n=161; 84.7%), primarily the YODA Project (n=157; 82.6%). All secondary publications published from 3 years before through the first 2 years after the primary publication (n=161) were internal (100%). Over time, however, external publications increased steadily, exceeding 50% of all secondary publications by year 11 and thereafter. External secondary publications were more frequently pooled analyses (151/209 [72.2%] vs 534/958 [55.7%]; *P*<0.001). Predictive or prognostic modelling (108/209 [51.7%] vs 322/958 [33.6%]; *P*<0.001), development of statistical models or algorithms (60/209 [28.7%] vs 114/958 [11.9%]; *P*<0.001), and validation of existing methods, models, or risk scores (32/209 [15.3%] vs 66/958 [6.9%]; *P*<0.001) were more frequent among external than internal secondary publications. Compared to internal secondary publications, external secondary publications were published in journals with higher impact factors (median, 6.7 [IQR, 3.4-16.6] vs 4.6 [2.9-10.2]; *P*=0.002) and had higher annual Altmetric Attention Scores (median, 2.1 [0.7-7.1] vs 0.6 [0.3-2.3]; *P*<0.001), but lower annual citation counts (median, 2.7 [1.1-5.6] vs 3.4 [1.6-7.5]; *P*<0.001) and were less likely to be cited in clinical guidelines (21/184 [11.4%] vs 235/805 [29.2%], *P*<0.001) or policy documents (14/184 [7.6%] vs 206/805 [25.6%], *P*<0.001); there was no difference in annual Mendeley reader counts (median, 7.4 [3.9-13.0] vs 8.0 [5.1-13.6], *P*=0.13).

**Conclusions:** Clinical trial data shared with external investigators through a data sharing platform generated substantial and sustained secondary research by both original trial investigators and external investigators. The proportion of secondary publications from any clinical trial generated by external investigators increased over time as external investigators pursued complementary research objectives that achieved a comparable scientific impact. Structured data sharing mechanisms may further enhance the scientific impact of clinical trials.

**What is already known on this topic:**

- Sharing individual participant-level data (IPD) from clinical trials can promote transparency, reproducibility, and secondary research.
- Several initiatives, including the Yale University Open Data Access (YODA) Project and government-supported data sharing platforms, provide external investigators with access to clinical trial data.
- While prior evaluations of secondary research generated from shared clinical trial data suggest that external investigators’ publications have citation impacts comparable to those of original trial investigators, overall evidence remains limited.

**What this study adds:**

- Analysis of 336 industry-sponsored clinical trials with IPD shared through the YODA Project showed that most generated secondary publications, by both original trial investigators and external investigators.
- The proportion of secondary publications from any clinical trial generated by external investigators increased over time, and compared with those generated by the original trial investigators, these publications more frequently use pooled analyses and focus on predictive or prognostic modelling and the development and validation of statistical methods.
- Secondary publications generated by external investigators were more often published in higher-impact journals and received higher Altmetric Attention Scores, but had lower annual citation counts and were less likely to be cited in clinical guidelines or policy documents than those generated by the original trial investigators.

## INTRODUCTION

Sharing individual participant-level data (IPD) from clinical trials promotes research transparency and reproducibility, builds public trust, and fosters innovation in medical research.^1^ Clinical trial data sharing typically involves providing external investigators with access to anonymized data through secure platforms managed by sponsors, academic institutions, or independent organizations.^2^ ^3^ Data may also be shared through direct collaborations or individual requests to the original trial investigators.^4–6^ Through access to clinical trial data collected by others, external investigators are able to examine new research questions, extending the scientific value of clinical trials beyond their original purpose and the contributions of trial participants,^7^ while potentially reducing redundant or duplicative research efforts and associated costs.^8^

While the value of clinical trial data sharing has been widely acknowledged,^9–12^ data sharing also presents important challenges.^13^ ^14^ Concerns persist around data misuse, breaches of participant confidentiality, protection of intellectual property, and the potential compromise of the original trial’s integrity due to invalid or selective secondary analyses.^15^ ^16^ In addition, questions remain about long-term impact and sustainability of data sharing. Establishing and maintaining responsible data sharing initiatives requires substantial investment in time, infrastructure, personnel, and administrative oversight.^12^ These efforts include building secure data repositories, processing and managing data access requests, and ensuring adherence to privacy and ethical standards, all of which require the ongoing commitment of resources. As a result, concerns have emerged regarding the long-term sustainability of such platforms and the scientific value generated by data sharing for stakeholders, including funding agencies, investigators, and the broader scientific community.

To date, there have been limited evaluations of the scientific impact generated through clinical trial data sharing initiatives. One study of the Digitalis Investigation Group (DIG) trial, whose IPD was made publicly available by the U.S. National Institutes of Health (NIH), found that more than half of the secondary publications using DIG trial data were generated by external investigators not affiliated with the original trial investigators, and their secondary publications had similar citation counts and were published in journals with comparable impact factors.^17^ Another study compared two distinct models of clinical trial data sharing—centralized versus decentralized repositories—used by two different NIH institutes and found centralized initiatives were associated with higher numbers of secondary publications, citation counts, journal h-index score, Altmetric Attention Scores, and abstract view counts.^18^

Additional and more comprehensive efforts are needed to characterize the scientific impact of secondary publications arising from shared clinical trial data and better understand the importance of these data sharing initiatives. Therefore, we examined the number, timing of publication, and scientific impact of secondary publications generated using data by both original trial investigators and external investigators from a portfolio of Johnson & Johnson-sponsored clinical trials shared through the Yale University Open Data Access (YODA) Project platform, based at Yale School of Medicine. This initiative, established over a decade ago, has made data from hundreds of Johnson & Johnson-sponsored clinical trials available to external investigators.^19^ ^20^

## METHODS

We conducted a cross-sectional study of secondary publications that used IPD from Johnson & Johnson-sponsored clinical trials listed on the YODA Project platform. This study did not require institutional review board approval or informed consent because it was based on publicly available information and involved no patient records, in accordance with 45 CFR §46. The study was prepared in accordance with the STROBE reporting guideline for cross-sectional studies.

### Trial Screening and Primary Publication Identification

We identified all Johnson & Johnson-sponsored clinical trials listed on the YODA Project platform with IPD available for external sharing as of December 31, 2021, allowing at least three years for investigators to use shared data and publish any resulting research, and with a full-length, peer-reviewed publication reporting results for the primary endpoint(s) by the original trial investigators (hereafter, the primary publication). We included only trials with an English-language primary publication.

Primary publications were identified through searches of Web of Science All Databases using the trial’s ClinicalTrials.gov identifier (NCT number). To ensure correct identification, we reviewed the introduction and methods sections of candidate publications to assess whether results had been previously reported and to confirm that the publication represented the first report of the trial’s primary endpoint(s). For trials for which a primary publication could not be identified using the NCT number in Web of Science, or for those without an NCT number, we conducted additional searches using the sponsor protocol number, trial name, or other trial-specific characteristics, including the generic and brand names of the study intervention, sample size, and key outcome measures or effect sizes reported for that trial. Trial characteristics used to identify the primary publications were extracted from clinical study report (CSRs) summaries available for trials on the YODA Project platform. Final confirmation of primary publications was obtained through direct communication with Johnson & Johnson.

### Secondary Publication Identification and Screening

Secondary publications were defined as peer-reviewed publications reporting original findings from analyses using IPD from one or more trials included in this study. We identified secondary publications by retrieving all citing records indexed in Web of Science All Databases for each primary publication. Searches were conducted between September 15 and December 15, 2024, and all records were updated through June 30, 2025.

All retrieved records were uploaded to Rayyan.ai, a web-based systematic review screening platform. Records were screened first at the title and abstract level. Full texts of potentially eligible records were reviewed to confirm use of IPD from one or more included trials. Publications were excluded if they were commentaries, editorials, letters to the editor, conference abstracts, or non-peer-reviewed papers. We also excluded studies that relied solely on CSRs or summary-level trial documents without analysis of trial IPD, as well as publications not written in English.

Screening was conducted by a team of five reviewers (ET, MM, HRH, SRM, and MF). To ensure consistency, records from the first 10 trials screened by each reviewer were independently validated through duplicate review by ET. After this initial validation phase, the remaining records were screened once, without duplication. Any uncertainties identified during screening were resolved through discussion with ET and JSR.

### Data Extraction and Classification

For all included trials and their corresponding primary publications, we extracted data into a structured spreadsheet (**Supplementary Data**). From the trials, we extracted product name, condition studied, trial phase, enrollment size, and the date the trial was made available for request on the YODA Project platform. From the primary publications, we extracted journal name, publication date, and author names. Included trials were classified by therapeutic area using the World Health Organization Anatomical Therapeutic Chemical (ATC) classification system. Classifications were restricted to ATC level 1, corresponding to the anatomical main group.

From each eligible secondary publication, we extracted publication date, journal name, author names and affiliations, and the reported data access mechanism, including whether IPD were obtained through a data sharing platform (such as the YODA Project), direct access from investigators or Johnson & Johnson, or other reported mechanisms. We also reviewed the content of each secondary publication to identify its primary research objective(s), which were categorized into one or more of the following: comparative effectiveness analyses; predictive or prognostic modelling; adverse event characterization and risk stratification; assessment of non-primary endpoint treatment effects; identification of mediators or moderators of treatment response; development of statistical models or algorithms; and validation of existing methods, models, or risk scores. Data extraction and classification were conducted by MM, HRH, SRM, and MF, with all records also independently extracted by ET. Any discrepancies were resolved through discussion among the reviewers.

#### Authorship Group

Secondary publications were classified as internal if they met either of the following criteria: (1) at least one author of the secondary publication was also an author on the primary publication or (2) at least one author was affiliated with Johnson & Johnson or one of its subsidiaries. Secondary publications using multiple trials were classified as internal if any of their authors were also authors on at least one of the primary publications associated with the trials included in the secondary publication. Secondary publications that did not meet either criterion were classified as external.

#### Analysis Type

Each secondary publication was categorized by type of analysis. Secondary analyses were studies that used IPD from a single trial. Pooled analyses involved combining IPD from two or more trials, encompassing but not limited to IPD meta-analyses. Studies that used IPD from more than one trial without combining IPD across trials were also classified as secondary analyses.

#### Scientific Impact Metrics

For each primary and secondary publication, we extracted journal impact factor (Journal Citation Reports), citation count (Web of Science All Databases), Altmetric Attention Score and Mendeley reader count (Altmetric Explorer), and recorded whether the publication was cited in clinical guidelines or policy documents (Altmetric Explorer). The Altmetric Attention Score is a measure of the level of online attention a publication receives across news media, social media, and other digital platforms, whereas the Mendeley reader count indicates the number of users who have saved the publication to their Mendeley reference libraries. To standardize impact metrics across different publication dates, we calculated yearly metrics by dividing each metric by the number of days since publication and multiplying by 365. Scientific impact metrics were extracted as of November 21, 2025.

#### Time From Primary Publication to Secondary Publication

We also calculated the time from primary to secondary publication in years by subtracting publication dates and dividing the number of days by 365. For secondary publications that included data from multiple trials, time differences were calculated between the publication date of the secondary publication and the publication date of each associated primary publication, and these differences were averaged to generate a single estimate per secondary publication.

### Statistical Analysis

We used descriptive statistics to summarize categorical variables as frequencies and relative frequencies. Continuous variables were summarized using medians and interquartile ranges (IQRs). Comparative analyses of journal impact factor, annual citation count, annual Altmetric Attention Score, and annual Mendeley reader count between internal and external secondary publications were performed using the Mann-Whitney U test. Differences in usage by clinical guidelines and policy documents, analysis type, research objectives, and therapeutic area between internal and external secondary publications were evaluated using the chi-square test. For the comparison of therapeutic area, the p-value was obtained using Monte Carlo simulation (10,000 replicates) to account for small, expected cell counts. A two-sided *P*<0.05 was considered statistically significant. All statistical analyses and visualizations were conducted using R version 4.4.2 (R Foundation for Statistical Computing, Vienna, Austria).

### Patient and public involvement

Patients and members of the public were not involved in the design, conduct, reporting, or dissemination plans of this study. This research analyzed publicly available information on a portfolio of clinical trials and their primary publications, as well as secondary publications using data from these trials. Although patients were not involved in this study, the clinical trial data used to generate these secondary publications could only have been possible because of the participatory efforts of patients. The sharing and reuse of clinical trial data aim to increase the scientific value of data generated through their participation. The findings of this study will contribute to ongoing discussions on clinical trial data sharing, with the goal of enhancing the value of these data to maximize the contributions made by trial participants.

## RESULTS

### Eligible Clinical Trials and Their Primary Publications

A total of 485 Johnson & Johnson-sponsored clinical trials listed on the YODA Project platform as of June 30, 2025, were assessed for eligibility. Of these, 336 trials (69.3%) met the eligibility criteria (**Figure 1**). The remaining 149 trials (30.7%) were excluded, including 93 (19.2%) that were made available on the YODA Project platform after December 31, 2021, 3 (0.6%) with a non-English-language primary publication, and 53 (10.9%) for which a primary publication could not be identified.

**Figure 1.**
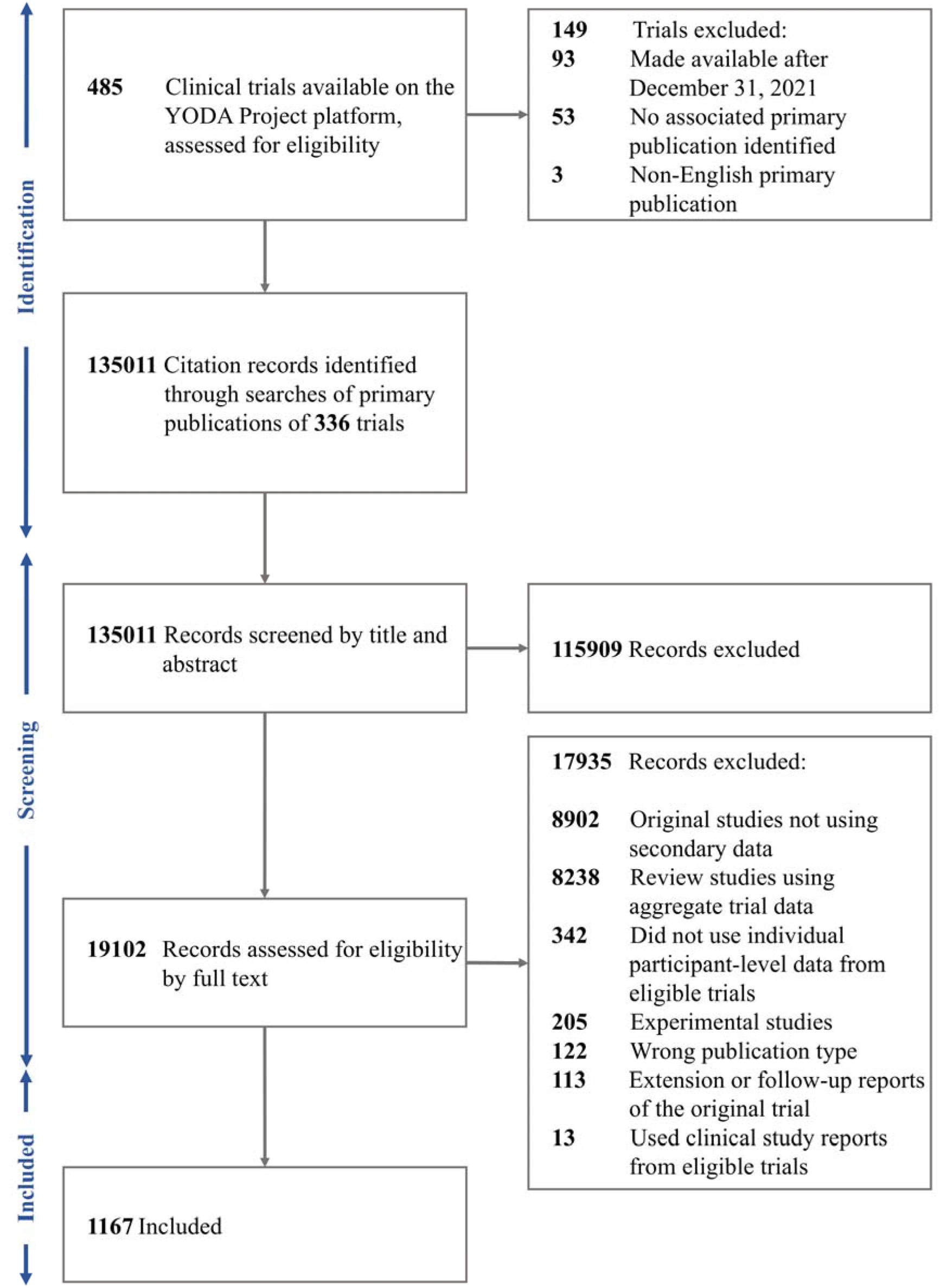
Flow Diagram for Identification of Eligible Clinical Trials and Screening of Secondary Publications Using Individual Participant-Level Data from These Trials

The 336 included trials evaluated a total of 37 medical products to treat 71 medical conditions. The complete lists of medical products and medical conditions represented within the included trials are provided in **Supplementary Tables 1 and 2**, respectively. Most trials were phase III (n=239; 71.1%) and enrolled more than 100 participants (n=279; 83.0%) (**Table 1**). The most frequently represented therapeutic area was the nervous system (n=160; 47.6%), followed by antineoplastic and immunomodulating agents (n=103; 30.7%). As of June 30, 2025, the median time that these trials had been available for request on the YODA Project platform was 7.8 years (IQR, 6.3-10.4).

**Table 1.** Characteristics of Johnson & Johnson-Sponsored Clinical Trials Available on the YODA Project Platform as of December 31, 2021, for Which an English-language Primary Publication Exists (n=336)

| Category | No. (%) |
| --- | --- |
| Trial phase |  |
| Phase I | 1 (0.3) |
| Phase I/II | 4 (1.2) |
| Phase II | 65 (19.3) |
| Phase II/III | 4 (1.2) |
| Phase III | 239 (71.1) |
| Phase IV | 21 (6.3) |
| No phase assigned | 2 (0.6) |
| Trial enrollment size |  |
| Median (interquartile range) | 309 (133-559) |
| Trials with ≤100 participants | 57 (17.0) |
| Trials with >100 participants | 279 (83.0) |
| Therapeutic Area |  |
| Alimentary tract and metabolism | 19 (5.7) |
| Anti-infectives for systemic use | 13 (3.9) |
| Antineoplastic and immunomodulating agents | 103 (30.7) |
| Nervous system | 160 (47.6) |
| Genitourinary system and sex hormones | 3 (0.9) |
| Cardiovascular system | 20 (6.0) |
| Blood and blood forming organs | 12 (3.6) |
| Antiparasitic products, insecticides, and repellents | 2 (0.6) |
| Dermatologicals | 2 (0.6) |
| Respiratory system | 2 (0.6) |

**Table 2.** Scientific Impact Metrics of Primary and Secondary Publications Associated with Johnson & Johnson-Sponsored Clinical Trials Available on the YODA Project Platform as of December 31, 2021.

| Impact Metrics | Primary Publications | Secondary Publications | Internal Secondary Publications | External Secondary Publications | P-value |
| --- | --- | --- | --- | --- | --- |
| Total, n (%) | 336 (100) | 1167 (100) | 958 (100) | 209 (100) |  |
| Median journal impact factor, No. (IQR) | 7.5<br>(3.8-21.0) | 4.7<br>(3.0-10.6) | 4.6<br>(2.9-10.2) | 6.7<br>(3.4-16.6) | 0.002<sup>a</sup> |
| Median annual citation, No. (IQR) count | 9.8<br>(4.1-28.5) | 3.3<br>(1.5-7.0) | 3.4<br>(1.6-7.5) | 2.7<br>(1.1-5.6) | <0.001<sup>a</sup> |
| Total, n (%)<sup>b</sup> | 322 (95.8) | 989 (84.7) | 805 (84.0) | 184 (88.0) |  |
| Usage by clinical guidelines, No. (%) | 233 (72.4) | 256 (25.9) | 235 (29.2) | 21 (11.4) | <0.001<sup>c</sup> |
| Usage by policy documents, No. (%) | 200 (62.1) | 220 (22.2) | 206 (25.6) | 14 (7.6) | <0.001<sup>c</sup> |
| Median annual Altmetric Attention Score, No. (IQR) | 0.9<br>(0.4-2.9) | 0.8<br>(0.3-2.9) | 0.6<br>(0.3-2.3) | 2.1<br>(0.7-7.1) | <0.001<sup>a</sup> |
| Median annual Mendeley reader count, No. (IQR) | 9.4<br>(5.3-16.9) | 7.9<br>(4.9-13.5) | 8.0<br>(5.1-13.6) | 7.4<br>(3.9-13.0) | 0.13<sup>a</sup> |

<sup>a</sup> P values were calculated using the Mann-Whitney U test for comparisons between internal and external secondary publications.
<sup>b</sup> Publications with available records in Altmetric Explorer.
<sup>c</sup> P values were calculated using the chi-square test for comparisons between internal and external secondary publications.

The primary publications associated with the included trials were published between 1992 and 2021. These publications appeared in journals with a median impact factor of 7.5 (IQR, 3.8-21.0) (**Table 2**). The median citation count was 152.5 (IQR, 66.8-437.5), corresponding to a median annual citation count of 9.8 (IQR, 4.1-28.5). Among the 336 primary publications, Altmetric Explorer indices were available for 322 (95.8%): 233 (72.4%) were cited in clinical guidelines (median, 2.0; IQR, 0.0-4.0), and 200 (62.1%) were cited in policy documents (median, 1.0; IQR, 0.0-4.0).

### Secondary Publications: Number and Timing of Publication

Citation searches of the primary publications associated with the 336 included trials identified 135,011 records in the Web of Science All Databases. Following title and abstract screening and subsequent full-text screening, 1,167 secondary publications met inclusion criteria. The reasons for exclusion at the full-text screening stage, along with a detailed description of the selection process, are shown in **Figure 1**.

Of the 336 included trials, 265 (78.9%) had at least one associated secondary publication, with a median of 3.0 secondary publications per trial (IQR, 1.0-10.0). Most secondary publications were internal publications (n=958; 82.1%). Overall, 238 trials (70.8%) had at least one associated internal secondary publication, with a median of 2.0 (IQR, 0.0-8.0) internal secondary publications per trial, whereas 152 (45.2%) trials had at least one associated external secondary publication, with a median of 0.0 (IQR, 0.0-3.0) external secondary publications per trial. 113 (33.6%) trials had only internal secondary publications, whereas 27 (8.0%) had only external secondary publications. The CANVAS trial (ClinicalTrials.gov identifier NCT01032629), which evaluated canagliflozin, had the highest number of associated secondary publications, with 103 in total, including 85 internal and 18 external publications (**Figure 2**).

**Figure 2.**
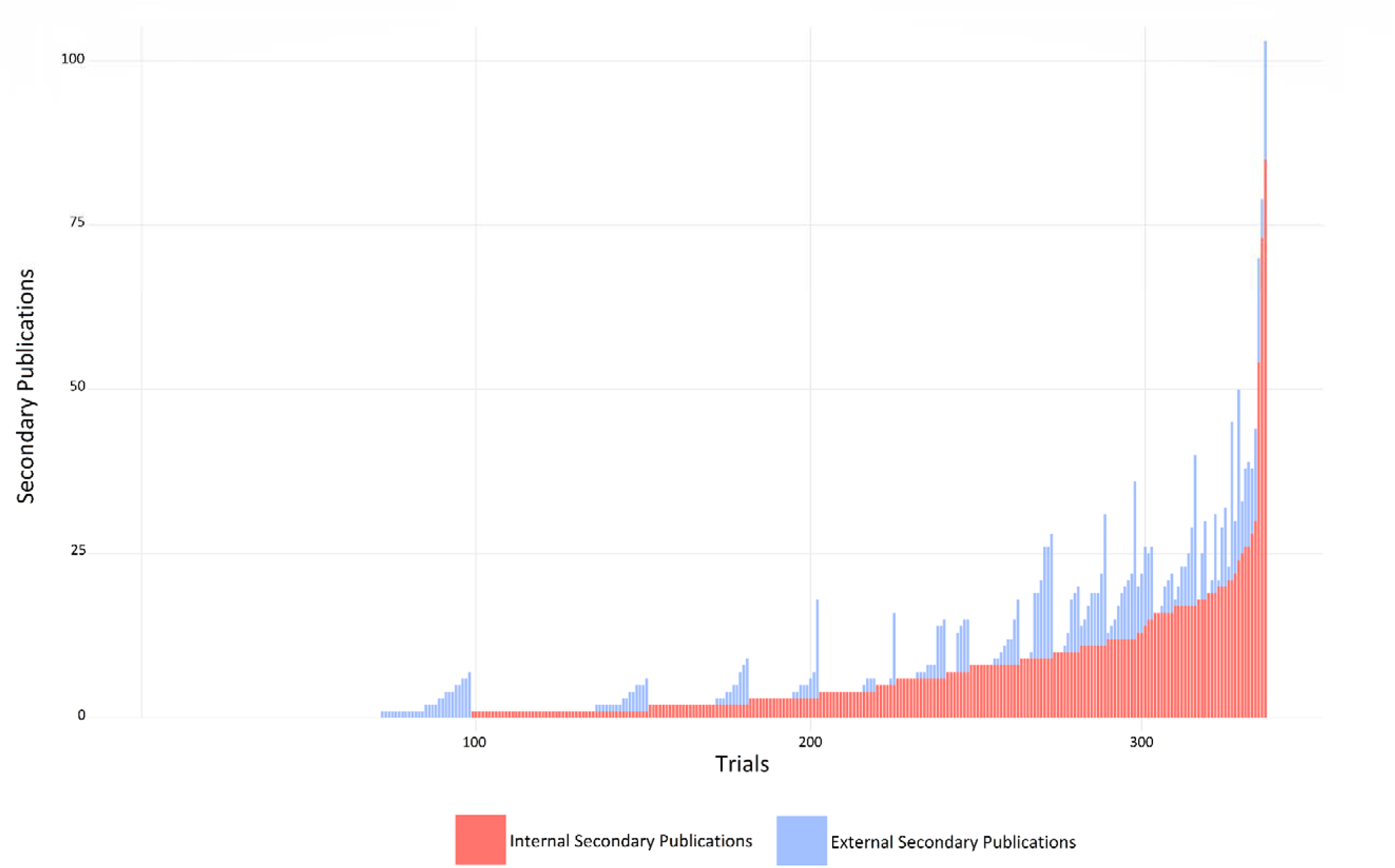
Distribution of Internal and External Secondary Publications Associated with Johnson & Johnson-Sponsored Clinical Trials Available on the YODA Project Platform as of December 31, 2021

Across all secondary publications, the median time from primary publication to secondary publication was 4.0 years (IQR, 2.3-6.6), with secondary publication timing ranging from 3 years before to 27 years after the primary publication. All 161 (100%) secondary publications from 3 years before through the first 2 years after the primary publication were internal. There was one instance of a secondary publication preceding the primary publication, a pooled analysis^21^ integrating data from four clinical trials, including the CANVAS trial, published in 2014, preceding publication of the CANVAS primary publication in 2017. Overall, internal secondary publications were published after significantly shorter periods of time than external secondary publications (median, 3.3 years [IQR, 1.9-5.3] vs 9.9 years [IQR, 6.5-13.5], *P*<0.001). The proportion of external secondary publications increased over time, exceeding 50% of all secondary publications by year 11 and thereafter (**Figure 3**).

**Figure 3.**
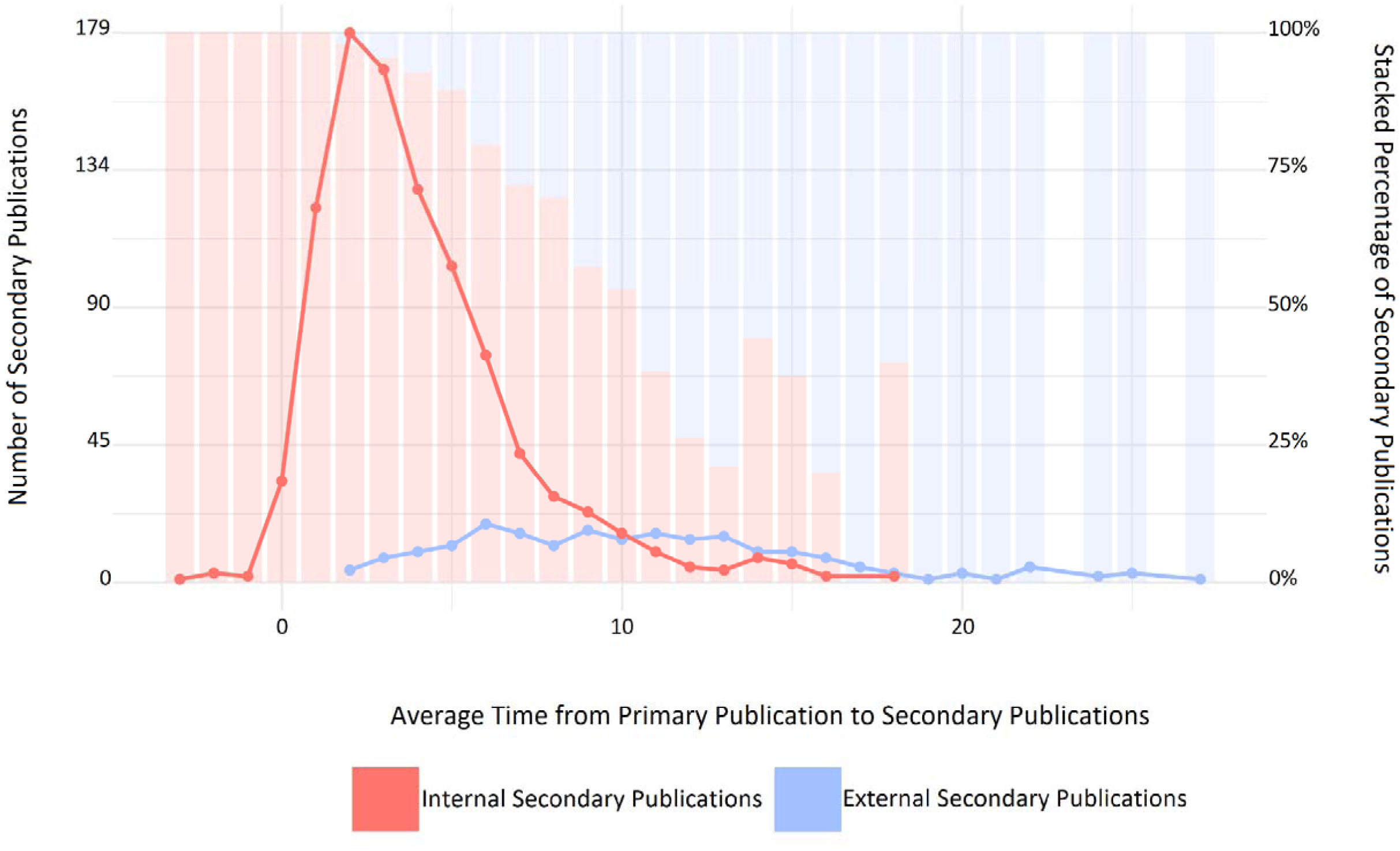
Time From Primary Publication to Internal and External Secondary Publications Associated with Johnson & Johnson-Sponsored Clinical Trials Available on the YODA Project Platform as of December 31, 2021 Note: For secondary publications associated with more than one trial, time from primary publication to secondary publication was calculated as the mean time difference between the publication date of the secondary publication and the publication dates of each associated primary publication. Time from primary publication to secondary publication was rounded to the nearest whole year in this Figure.

Among external secondary publications for which the data access mechanism was reported (190 of 209; 90.9%), most obtained access through data sharing platforms (n=161; 84.7%), primarily the YODA Project (n=157; 82.6%). Other external secondary publications accessed data directly from investigators or from Johnson & Johnson (n=20; 10.5%) or were conducted by the U.S. Food and Drug Administration (FDA) using its own databases of trials previously submitted as part of the evidence for medical product application review (n=9; 4.7%).

### Secondary Publications: Characteristics

Most secondary publications were pooled analyses (n=685; 58.7%), which were more common among external than internal secondary publications (151 of 209 [72.2%] vs 534 of 958 [55.7%], *P*<0.001) (**Table 3**). The distribution of secondary publications across therapeutic areas differed significantly between internal and external secondary publications (*P*<0.001), with a higher proportion of internal secondary publications involving trials of the alimentary tract and metabolism (175 of 958 [18.3%] vs 21 of 209 [10.0%]) and a higher proportion of external secondary publications involving trials of multiple therapeutic areas (13 of 209 [6.2%] vs 3 of 958 [0.3%]).

**Table 3.**
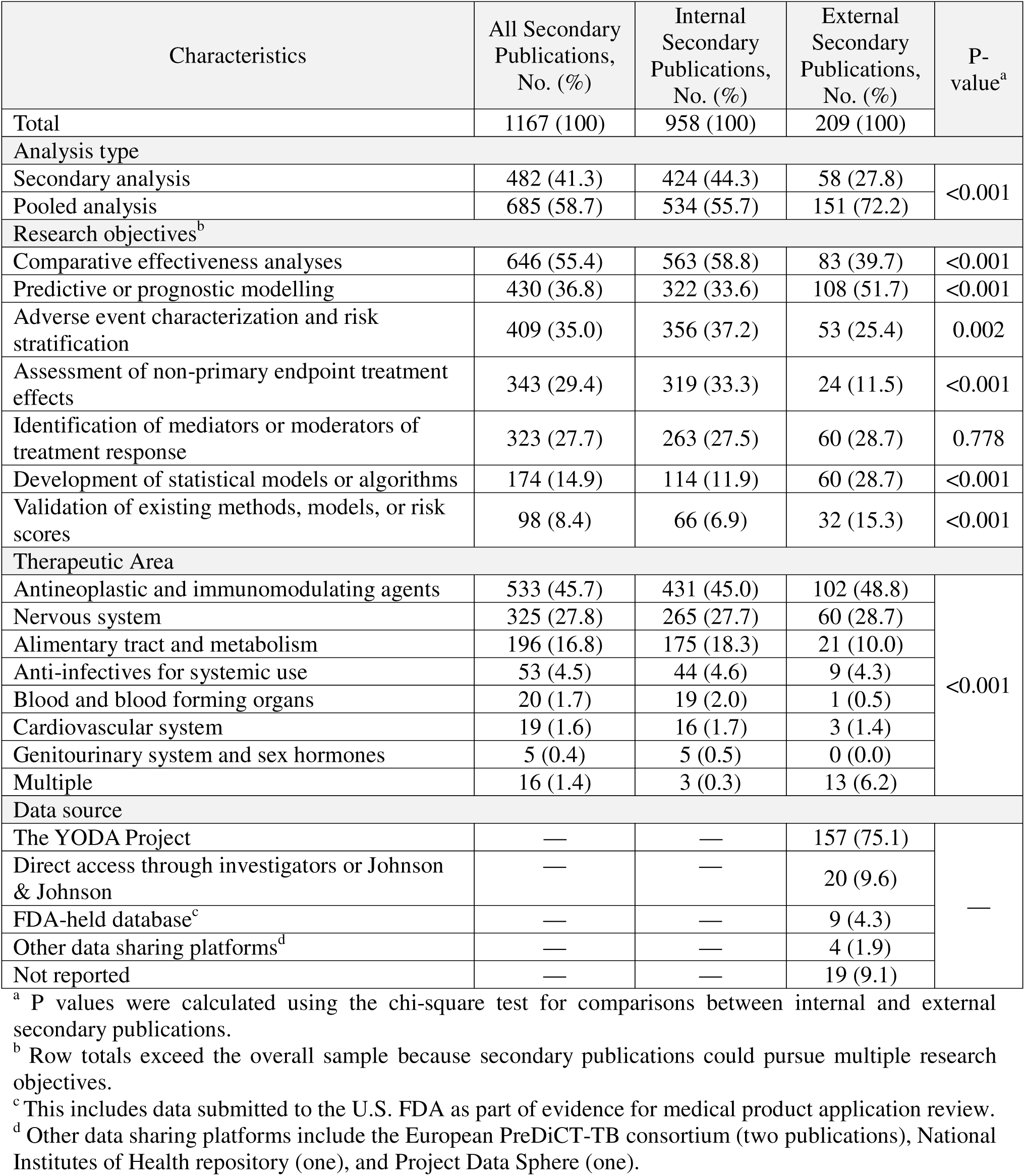
Characteristics of Secondary Publications Associated with Johnson & Johnson-Sponsored Clinical Trials Available on the YODA Project Platform as of December 31, 2021.

| Characteristics | All Secondary Publications, No. (%) | Internal Secondary Publications, No. (%) | External Secondary Publications, No. (%) | P-value<sup>a</sup> |
| --- | --- | --- | --- | --- |
| Total | 1167 (100) | 958 (100) | 209 (100) |  |
| Analysis type |  |  |  |  |
| Secondary analysis | 482 (41.3) | 424 (44.3) | 58 (27.8) | <0.001 |
| Pooled analysis | 685 (58.7) | 534 (55.7) | 151 (72.2) |  |
| Research objectives<sup>b</sup> |  |  |  |  |
| Comparative effectiveness analyses | 646 (55.4) | 563 (58.8) | 83 (39.7) | <0.001 |
| Predictive or prognostic modelling | 430 (36.8) | 322 (33.6) | 108 (51.7) | <0.001 |
| Adverse event characterization and risk stratification | 409 (35.0) | 356 (37.2) | 53 (25.4) | 0.002 |
| Assessment of non-primary endpoint treatment effects | 343 (29.4) | 319 (33.3) | 24 (11.5) | <0.001 |
| Identification of mediators or moderators of treatment response | 323 (27.7) | 263 (27.5) | 60 (28.7) | 0.778 |
| Development of statistical models or algorithms | 174 (14.9) | 114 (11.9) | 60 (28.7) | <0.001 |
| Validation of existing methods, models, or risk scores | 98 (8.4) | 66 (6.9) | 32 (15.3) | <0.001 |
| Therapeutic Area |  |  |  |  |
| Antineoplastic and immunomodulating agents | 533 (45.7) | 431 (45.0) | 102 (48.8) | <0.001 |
| Nervous system | 325 (27.8) | 265 (27.7) | 60 (28.7) |  |
| Alimentary tract and metabolism | 196 (16.8) | 175 (18.3) | 21 (10.0) |  |
| Anti-infectives for systemic use | 53 (4.5) | 44 (4.6) | 9 (4.3) |  |
| Blood and blood forming organs | 20 (1.7) | 19 (2.0) | 1 (0.5) |  |
| Cardiovascular system | 19 (1.6) | 16 (1.7) | 3 (1.4) |  |
| Genitourinary system and sex hormones | 5 (0.4) | 5 (0.5) | 0 (0.0) |  |
| Multiple | 16 (1.4) | 3 (0.3) | 13 (6.2) |  |
| Data source |  |  |  |  |
| The YODA Project | — | — | 157 (75.1) | — |
| Direct access through investigators or Johnson & Johnson | — | — | 20 (9.6) |  |
| FDA-held database<sup>c</sup> | — | — | 9 (4.3) |  |
| Other data sharing platforms<sup>d</sup> | — | — | 4 (1.9) |  |
| Not reported | — | — | 19 (9.1) |  |

<sup>a</sup> P values were calculated using the chi-square test for comparisons between internal and external secondary publications.
<sup>b</sup> Row totals exceed the overall sample because secondary publications could pursue multiple research objectives.
<sup>c</sup> This includes data submitted to the U.S. FDA as part of evidence for medical product application review.
<sup>d</sup> Other data sharing platforms include the European PreDiCT-TB consortium (two publications), National Institutes of Health repository (one), and Project Data Sphere (one).

Across all 1,167 secondary publications, the most common research objectives were comparative effectiveness analyses (n=646; 55.4%) and predictive or prognostic modelling (n=430; 36.8%). Predictive or prognostic modelling (108 of 209 [51.7%] vs 322 of 958 [33.6%], *P*<0.001), development of statistical models or algorithms (60 of 209 [28.7%] vs 114 of 958 [11.9%], *P*<0.001), and validation of existing methods, models, or risk scores (32 of 209 [15.3%] vs 66 of 958 [6.9%], *P*<0.001) were more frequent among external secondary publications than internal secondary publications. In contrast, comparative effectiveness analyses (563 of 958 [58.8%] vs 83 of 209 [39.7%], *P*<0.001), adverse event characterization and risk stratification (356 of 958 [37.2%] vs 53 of 209 [25.4%], *P*=0.002), and assessments of non-primary endpoint treatment effects (319 of 958 [33.3%] vs 24 of 209 [11.5%], *P*<0.001) were more common among internal secondary publications than external secondary publications.

### Secondary Publications: Scientific Impact Metrics

External secondary publications were published in journals with higher impact factors when compared with internal secondary publications (median, 6.7 [IQR, 3.4-16.6] vs 4.6 [IQR, 2.9-10.2], *P*=0.002) and had higher annual Altmetric Attention Scores (median, 2.1 [IQR, 0.7-7.1] vs 0.6 [IQR, 0.3-2.3], *P*<0.001) (**Table 2**). In contrast, annual citation counts were lower for external than for internal secondary publications (median, 2.7 [IQR, 1.1-5.6] vs 3.4 [IQR, 1.6-7.5], *P*<0.001). There was no difference in annual Mendeley reader counts between external and internal secondary publications (median, 7.4 [IQR, 3.9-13.0] vs 8.0 [IQR, 5.1-13.6], *P*=0.13).

Among secondary publications with available Altmetric Explorer indices (989 of 1,167; 84.7%), external secondary publications were less likely than internal secondary publications to be cited in clinical guidelines (21 of 184 [11.4%] vs 235 of 805 [29.2%], *P*<0.001) or policy documents (14 of 184 [7.6%] vs 206 of 805 [25.6%], *P*<0.001).

## DISCUSSION

In this cross-sectional study of a large portfolio of Johnson & Johnson-sponsored clinical trials shared with external investigators through the YODA Project platform, we found that data were frequently used by both original trial investigators and external investigators unaffiliated with the original trial teams to generate a large body of research, including numerous secondary publications over time. Although secondary publications by original trial investigators (i.e., internal secondary publications) accounted for a larger share overall, the proportion of secondary publications from any clinical trial generated by external investigators (i.e., external secondary publications) increased over time. Moreover, external investigators pursued complementary research objectives that achieved a comparable scientific impact, using pooled analyses for predictive or prognostic modelling and development or validation of statistical models. These findings suggest that structured data sharing mechanisms may further enhance the scientific impact of clinical trials.

Internal secondary publications predominated in the years immediately following the primary publication of the clinical trial, reflecting continued analyses by the original trial investigators, often of important secondary endpoints. In contrast, external secondary publications were generated later and increased steadily in subsequent years, becoming a larger proportion of the overall number of secondary publications after more than a decade. Similar temporal patterns have been observed in prior evaluations of shared clinical trial data, including analyses of the DIG trial, in which publications by external investigators continued after publications by the original trial investigators declined.^17^ These findings suggest that shared clinical trial data can continue to generate new insights and knowledge many years after primary publication, with external investigators extending the scientific lifespan of trials beyond the period of active involvement with the data by the original trial investigators.

The research objectives of secondary publications differed between those conducted by original trial investigators and external investigators, suggesting complementary patterns of clinical trial data use. Internal secondary publications more frequently evaluated comparative effectiveness, non-primary endpoint treatment effects, and adverse event characterization and risk stratification, further characterizing the efficacy and safety of the medical products studied in the trials. In contrast, external secondary publications more frequently focused on predictive or prognostic modelling, as well as the development and validation of statistical methods, consistent with findings from prior evaluations of data requests to major clinical data sharing platforms.^3^ These differences underscore the importance of data sharing, which enables clinical trial data to be used not only to further characterize treatment effects but also to support methodological innovation and risk prediction beyond the original objectives of the trials and beyond the analyses conducted and reported by the original trial investigators.

External secondary publications achieved a comparable scientific impact in relation to internal secondary publications. For instance, external secondary publications were published in journals with higher impact factors than internal secondary publications and received higher Altmetric Attention Scores, which may reflect their greater use of pooled analyses and the resulting larger study populations, increased statistical power, and more precise effect estimates, features that might raise the priority of the research and confidence in its findings for higher-impact journals.^22^ However, external secondary publications had lower annual citation counts and were cited less frequently in clinical guidelines and policy documents than internal secondary publications. While these differences may partly reflect variation in research objectives, it is also possible that because internal secondary publications had been published earlier than external secondary publications, they had more opportunity to accrue the attention needed to be cited by peer scientists and by professional societies for clinical guidelines and policy documents, even despite our attempts to standardize metrics using annual citation counts.^23^ ^24^ Nevertheless, the small observed differences between internal and external secondary publications suggest broadly similar levels of scholarly engagement and scientific impact. Independent of these comparisons, external secondary publications, contributed to the clinical, policy, and broader scientific literature, contributions that would not have occurred in the absence of data sharing.

Most external secondary publications relied on formal data sharing platforms for access to the clinical trial data, most commonly through the YODA Project, rather than direct access through the original trial investigators or sponsors, highlighting the importance of structured data sharing mechanisms that facilitate independent secondary research. Over the past decade, regulatory bodies, funding agencies, academic institutions, and industry sponsors have implemented policies to support responsible sharing of clinical trial data.^8^ ^25–33^ For example, the National Institute for Health and Care Research has introduced policies encouraging data sharing, including requirements for data management plans and data availability statements for funded research, and has supported secure research infrastructures such as the Clinical Practice Research Datalink, which allow external investigators to access anonymized health data within controlled environments. In parallel, initiatives such as the YODA Project and Vivli have created structured data sharing mechanisms through which external investigators can request access to IPD from clinical trials. Despite these efforts, access to clinical trial data remains incomplete, and many trials still do not make IPD and supporting documentation available through formal data sharing mechanisms.^34^ ^35^ Continued policy support, further development of transparent and sustainable data sharing infrastructures, and promotion of a broader culture of responsible data sharing will therefore be important to maximize the scientific value of clinical trials and enable broader independent evaluation and reuse of trial data.

Several limitations of this study should be taken into consideration. First, the study was restricted to clinical trials sponsored by one large medical product company, Johnson & Johnson, which may limit the generalizability of findings to other clinical trial portfolios, such as those conducted in different therapeutic areas or involving different medical products and conditions, although the range of medical products and conditions studied among the sample of 336 trials was broad. Second, secondary publications were identified through citation screening of primary publications in Web of Science as of June 30, 2025. Although we screened a large number of records, totaling more than 135,000, secondary publications that did not cite the primary publication, or did so but were published after July 1, 2025, may not have been identified, potentially leading to underestimation of secondary publications, which, based on our findings, would be more likely to affect estimates of external secondary publications. Third, because of the large volume of records, screening was conducted by a single reviewer per record after initial validation through duplicate screening, raising the possibility that some eligible secondary publications were missed. Fourth, scientific impact was assessed with readily available measures but may not capture all aspects of value prioritized by the scientific enterprise. Finally, this study did not have a publicly registered protocol; however, a protocol was developed within the study team before screening of the clinical trials listed on the YODA Project platform was initiated.

## CONCLUSIONS

Clinical trial data, shared through the YODA Project platform, generated substantial and sustained secondary research by both original trial investigators and external investigators. The proportion of secondary publications from any clinical trial generated by external investigators increased over time as external investigators pursued complementary research objectives that achieved a comparable scientific impact. Structured data sharing mechanisms may further enhance the scientific impact of clinical trials. Importantly, sharing clinical trial data offers benefits beyond measures of scientific impact, such as enabling independent analyses, promoting transparency, reducing the patient and sponsor resources that might otherwise be spent on the conduct of potentially duplicative trials, and supporting the broader medical research enterprise. Continued policy support, development of sustainable data sharing infrastructure, and promotion of a broader culture of responsible data sharing will be important to increase the long-term scientific value of clinical trial data.

## Ethics statements

## Ethical approval

This study did not require institutional review board approval or informed consent because it was based on publicly available information and involved no patient records, in accordance with 45 CFR §46.

## Data availability statement

All data supporting the findings of this study are available in the Supplementary Data provided with this article.

## Acknowledgments

None.

## Contributors

ET, HRH, JAB, NRD, CPG, RL, JDR, JDW, MKW, HMK, and JSR conceived and designed the study. ET, SB, KC, DH, JDR, TS, JW, and JSR were involved in trial screening and identification of primary publications. ET, MM, HRH, SRM, and MF conducted secondary publication identification and screening and performed data extraction. ET curated the data and conducted the descriptive analyses and visualizations. All authors contributed to interpretation of the findings. ET drafted the initial manuscript. All authors critically revised the manuscript and approved the final version for submission. JSR supervised the study and is the guarantor. The corresponding author attests that all listed authors meet authorship criteria and that no others meeting the criteria have been omitted.

## Transparency

The corresponding author (JSR) affirms that the manuscript is an honest, accurate, and transparent account of the study being reported; that no important aspects of the study have been omitted; and that any discrepancies from the study as registered have been explained.

## Dissemination to participants and related patient and public communities

Results from this study will be disseminated through social media platforms (X, LinkedIn, and Bluesky) and will be shared with relevant stakeholders. As no patients or members of the public were recruited or involved in this study, there are no plans to disseminate the results directly to any patients or members of the public.

## REFERENCES

1. Institute of M. The National Academies Collection: Reports funded by National Institutes of Health. Sharing Clinical Research Data: Workshop Summary. Washington (DC): National Academies Press (US) Copyright © 2013 by the National Academy of Sciences. All rights reserved. 2013.

2. Ohmann C, Banzi R, Canham S, et al. Sharing and reuse of individual participant data from clinical trials: principles and recommendations. BMJ Open 2017;7(12):e018647. doi: 10.1136/bmjopen-2017-018647 [published Online First: 20171214]

3. Vazquez E, Gouraud H, Naudet F, et al. Characteristics of available studies and dissemination of research using major clinical data sharing platforms. Clin Trials 2021;18(6):657–66. doi: 10.1177/17407745211038524 [published Online First: 20210818]

4. Rising K, Bacchetti P, Bero L. Reporting bias in drug trials submitted to the Food and Drug Administration: review of publication and presentation. PLoS Med 2008;5(11):e217; discussion e17. doi: 10.1371/journal.pmed.0050217

5. Ross JS, Mulvey GK, Hines EM, et al. Trial publication after registration in ClinicalTrials.Gov: a cross-sectional analysis. PLoS Med 2009;6(9):e1000144. doi: 10.1371/journal.pmed.1000144 [published Online First: 20090908]

6. Chan AW, Hróbjartsson A, Haahr MT, et al. Empirical evidence for selective reporting of outcomes in randomized trials: comparison of protocols to published articles. Jama 2004;291(20):2457–65. doi: 10.1001/jama.291.20.2457

7. Chan AW, Song F, Vickers A, et al. Increasing value and reducing waste: addressing inaccessible research. Lancet 2014;383(9913):257–66. doi: 10.1016/s0140-6736(13)62296-5 [published Online First: 20140108]

8. Ross JS, Krumholz HM. Ushering in a new era of open science through data sharing: the wall must come down. Jama 2013;309(13):1355–6. doi: 10.1001/jama.2013.1299

9. Hudson KL, Collins FS. Sharing and reporting the results of clinical trials. Jama 2015;313(4):355–6. doi: 10.1001/jama.2014.10716

10. Kiley R, Peatfield T, Hansen J, et al. Data Sharing from Clinical Trials - A Research Funder’s Perspective. N Engl J Med 2017;377(20):1990–92. doi: 10.1056/NEJMsb1708278

11. Modjarrad K, Moorthy VS, Millett P, et al. Developing Global Norms for Sharing Data and Results during Public Health Emergencies. PLoS Med 2016;13(1):e1001935. doi: 10.1371/journal.pmed.1001935 [published Online First: 20160105]

12. Committee on Strategies for Responsible Sharing of Clinical Trial D, Board on Health Sciences P, Institute of M. Sharing Clinical Trial Data: Maximizing Benefits, Minimizing Risk. Washington (DC): National Academies Press (US) Copyright 2015 by the National Academy of Sciences. All rights reserved. 2015.

13. Mello MM, Lieou V, Goodman SN. Clinical Trial Participants’ Views of the Risks and Benefits of Data Sharing. New England Journal of Medicine 2018;378(23):2202–11. doi: doi:10.1056/NEJMsa1713258

14. Devereaux PJ, Guyatt G, Gerstein H, et al. Toward Fairness in Data Sharing. N Engl J Med 2016;375(5):405–7. doi: 10.1056/NEJMp1605654

15. National Academies of Sciences E, Medicine, Health, et al. The National Academies Collection: Reports funded by National Institutes of Health. In: Shore C, Hinners J, Khandekar E, et al., eds. Reflections on Sharing Clinical Trial Data: Challenges and a Way Forward: Proceedings of a Workshop. Washington (DC): National Academies Press (US) Copyright 2020 by the National Academy of Sciences. All rights reserved. 2020.

16. Ross JS, Lehman R, Gross CP. The importance of clinical trial data sharing: toward more open science. Circ Cardiovasc Qual Outcomes 2012;5(2):238–40. doi: 10.1161/circoutcomes.112.965798

17. Angraal S, Ross JS, Dhruva SS, et al. Merits of Data Sharing: The Digitalis Investigation Group Trial. J Am Coll Cardiol 2017;70(14):1825–27. doi: 10.1016/j.jacc.2017.07.786

18. Rowhani-Farid A, Grewal M, Solar S, et al. Clinical trial data sharing: a cross-sectional study of outcomes associated with two U.S. National Institutes of Health models. Sci Data 2023;10(1):529. doi: 10.1038/s41597-023-02436-0 [published Online First: 20230808]

19. Ross JS, Waldstreicher J, Bamford S, et al. Overview and experience of the YODA Project with clinical trial data sharing after 5 years. Sci Data 2018;5:180268. doi: 10.1038/sdata.2018.268 [published Online First: 20181127]

20. Taherifard E, Ritchie JD, Willeford MK, et al. A Decade of the YODA Project: Advancing Clinical Trial Data Sharing through Transparency, Independence, and Collaboration. Under review., 2025.

21. Yamout H, Perkovic V, Davies M, et al. Efficacy and safety of canagliflozin in patients with type 2 diabetes and stage 3 nephropathy. Am J Nephrol 2014;40(1):64–74. doi: 10.1159/000364909 [published Online First: 20140717]

22. Heidenreich A, Eisemann N, Katalinic A, et al. Study results from journals with a higher impact factor are closer to “truth“: a meta-epidemiological study. Syst Rev 2023;12(1):8. doi: 10.1186/s13643-023-02167-8 [published Online First: 20230118]

23. Pérez-Campdesuñer R, Sánchez-Rodríguez A, Martínez-Vivar R, et al. Bias in Citation Visibility: Temporal Dynamics and the Unequal Life Cycle of Academic Articles—Evidence from SME and Internationalization Research. Publications 2025;13(4):62.

24. Aksnes DW. Characteristics of highly cited papers. Research Evaluation 2003;12(3):159–70. doi: 10.3152/147154403781776645

25. Rydzewska LHM, Stewart LA, Tierney JF. Sharing individual participant data: through a systematic reviewer lens. Trials 2022;23(1):167. doi: 10.1186/s13063-021-05787-4

26. DeVito NJ, French L, Goldacre B. Noncommercial Funders’ Policies on Trial Registration, Access to Summary Results, and Individual Patient Data Availability. Jama 2018;319(16):1721–23. doi: 10.1001/jama.2018.2841

27. Goldacre B, Lane S, Mahtani KR, et al. Pharmaceutical companies’ policies on access to trial data, results, and methods: audit study. Bmj 2017;358:j3334. doi: 10.1136/bmj.j3334 [published Online First: 20170726]

28. Miller J, Ross JS, Wilenzick M, et al. Sharing of clinical trial data and results reporting practices among large pharmaceutical companies: cross sectional descriptive study and pilot of a tool to improve company practices. Bmj 2019;366:l4217. doi: 10.1136/bmj.l4217 [published Online First: 20190710]

29. Shaw DL, Ross JS. US Federal Government Efforts to Improve Clinical Trial Transparency with Expanded Trial Registries and Open Data Sharing. AMA J Ethics 2015;17(12):1152–9. doi: 10.1001/journalofethics.2015.17.12.pfor1-1512 [published Online First: 20151201]

30. Johnson AL, Anderson JM, Bouvette M, et al. Clinical trial data-sharing policies among journals, funding agencies, foundations, and other professional organizations: a scoping review. Journal of Clinical Epidemiology 2023;154:42–55. doi: 10.1016/j.jclinepi.2022.11.009

31. Ross JS, Waldstreicher J, Krumholz HM. Data Sharing - A New Era for Research Funded by the U.S. Government. N Engl J Med 2023;389(26):2408–10. doi: 10.1056/NEJMp2308792 [published Online First: 20231115]

32. Tannenbaum S, Ross JS, Krumholz HM, et al. Early Experiences With Journal Data Sharing Policies: A Survey of Published Clinical Trial Investigators. Ann Intern Med 2018;169(8):586–88. doi: 10.7326/m18-0723 [published Online First: 20180717]

33. Gonzales S, Carson MB, Holmes K. Ten simple rules for maximizing the recommendations of the NIH data management and sharing plan. PLoS Comput Biol 2022;18(8):e1010397. doi: 10.1371/journal.pcbi.1010397 [published Online First: 20220803]

34. Chen H, Zhao Y, Cao B, et al. Breaking the silence of sharing data in medical research. PLoS One 2024;19(5):e0301917. doi: 10.1371/journal.pone.0301917 [published Online First: 20240529]

35. Ohmann C, Moher D, Siebert M, et al. Status, use and impact of sharing individual participant data from clinical trials: a scoping review. BMJ Open 2021;11(8):e049228. doi: 10.1136/bmjopen-2021-049228 [published Online First: 20210818]

