## Supplement for "Assessing the Secondary Use and Scientific Impact of Shared Clinical Trial Data: A Cross-Sectional Study of Clinical Trials Shared on the YODA Project Platform"

**Running title:** Scientific Impact of Clinical Trial Data Sharing

Erfan Taherifard^1^, Maryam Mooghali^1^, Hollin R. Hakimian^2^, Sahil R. Mane^3^, Mengyuan Fu^1^, Stephen Bamford^4^, Jesse A. Berlin^5^, Karla Childers^6^, Nihar R. Desai^7^, Cary P. Gross^1,8,9^, Debbie Hewens^4^, Richard Lehman^10^, Jessica D. Ritchie^1^, Tamsin Sargood^4^, Joanne Waldstreicher^11^, Joshua D. Wallach^12^, Molly K. Willeford^1^, Harlan M. Krumholz^7,13,14^, Joseph S. Ross^1,8,13,14^

1. Section of General Internal Medicine, Yale School of Medicine, New Haven, CT, USA
2. Learning Design and Technology, Graduate School of Education, Stanford University, Stanford, CA, USA
3. Washington University School of Medicine in St. Louis, St. Louis, MO, USA
4. Johnson & Johnson, England, UK
5. Center for Pharmacoepidemiology and Treatment Science, Department of Biostatistics and Epidemiology, School of Public Health, Rutgers University, Piscataway, NJ, USA
6. Johnson & Johnson, New Brunswick, NJ, USA
7. Section of Cardiovascular Medicine, Yale School of Medicine, New Haven, CT, USA
8. National Clinician Scholars Program, Yale School of Medicine, New Haven, CT, USA
9. Department of Chronic Disease Epidemiology, Yale School of Public Health, New Haven, CT, USA
10. Institute of Applied Health Research, University of Birmingham, England, UK
11. Johnson & Johnson (retired), New Brunswick, NJ, USA.
12. Department of Epidemiology, Rollins School of Public Health, Emory University, Atlanta, GA, USA
13. Yale-New Haven Hospital Center for Outcomes Research and Evaluation, New Haven, CT, USA
14. Department of Health Policy and Management, Yale School of Public Health, New Haven, CT, USA

**Corresponding author:**

Joseph S. Ross, MD, MHS, Professor of Medicine and Public Health; Co-Director, Yale Open Data Access (YODA) Project, Center for Outcomes Research and Evaluation (CORE), 195 Church Street, 6th Floor, New Haven, CT 06510, USA;

Table of Contents

Supplementary Table 1	3

Supplementary Table 2	4

Supplementary Table 1. Medical Products Evaluated in Included Johnson & Johnson-Sponsored Clinical Trials Available on the YODA Project Platform as of December 31, 2021

| Products | No. (%) |
| --- | --- |
| TOPAMAX® | 37 (11.0) |
| RISPERDAL® | 25 (7.4) |
| INVEGA® | 23 (6.8) |
| STELARA® | 22 (6.5) |
| SIMPONI® | 20 (6.0) |
| INVEGA® SUSTENNA® | 18 (5.4) |
| REMICADE® | 18 (5.4) |
| INVOKANA® | 18 (5.4) |
| RAZADYNE® | 16 (4.8) |
| RISPERDAL® CONSTA® | 16 (4.8) |
| ZYTIGA® | 16 (4.8) |
| TRACLEER® | 13 (3.9) |
| CONCERTA® | 12 (3.6) |
| SPRAVATO® | 12 (3.6) |
| PROCRIT® | 9 (2.7) |
| YONDELIS® | 8 (2.4) |
| OPSUMIT® | 6 (1.8) |
| PREZISTA® | 6 (1.8) |
| DARZALEX® | 6 (1.8) |
| PLIVENSIA™ | 4 (1.2) |
| IMBRUVICA® | 3 (0.9) |
| MONONESSA ® ORTHO-CYCLEN ® ORTHO TRI-CYCLEN ® TRINESSA ® | 3 (0.9) |
| SIRTURO® | 3 (0.9) |
| UPTRAVI® | 3 (0.9) |
| EDURANT® | 2 (0.6) |
| Bapineuzumab | 2 (0.6) |
| ROGAINE® | 2 (0.6) |
| JNJ-38518168 | 2 (0.6) |
| TREMFYA® | 2 (0.6) |
| VERMOX® | 2 (0.6) |
| DOXIL® | 1 (0.3) |
| ERLEADA® | 1 (0.3) |
| INTELENCE® | 1 (0.3) |
| LEVAQUIN® | 1 (0.3) |
| Razadyne®/Reminyl | 1 (0.3) |
| THERMOCOOL® SMARTTOUCH™ Catheter | 1 (0.3) |
| ZAVESCA® | 1 (0.3) |

**Supplementary Table 2.** Medical Conditions Evaluated in Included Johnson & Johnson-Sponsored Clinical Trials Available on the YODA Project Platform as of December 31, 2021

| Medical Conditions | No. (%) |
| --- | --- |
| Schizophrenia | 51 (15.2) |
| Alzheimer's disease | 20 (6.0) |
| Bipolar disorder | 19 (5.7) |
| Diabetes mellitus, type 2 | 17 (5.1) |
| Rheumatoid arthritis | 17 (5.1) |
| Prostatic neoplasm | 16 (4.8) |
| Major depressive disorder | 15 (4.5) |
| Psoriasis | 15 (4.5) |
| Attention-deficit/hyperactivity disorder | 13 (3.9) |
| Crohn's disease | 12 (3.6) |
| Obesity | 11 (3.3) |
| Epilepsy | 9 (2.7) |
| HIV infection | 9 (2.7) |
| Migraine disorders | 8 (2.4) |
| Pulmonary arterial hypertension | 8 (2.4) |
| Ulcerative colitis | 8 (2.4) |
| Multiple myeloma | 6 (1.8) |
| Ankylosing spondylitis | 5 (1.5) |
| Attention deficit and disruptive behavior disorder | 4 (1.2) |
| Dementia | 4 (1.2) |
| Digital ulcers in systemic sclerosis | 4 (1.2) |
| Pulmonary hypertension | 4 (1.2) |
| Sarcoma | 4 (1.2) |
| Anemia | 3 (0.9) |
| Autistic disorder | 3 (0.9) |
| Idiopathic pulmonary fibrosis | 3 (0.9) |
| Migraine | 3 (0.9) |
| Schizoaffective disorder | 3 (0.9) |
| Tuberculosis | 3 (0.9) |
| Androgenetic alopecia | 2 (0.6) |
| Anemia in critical illness | 2 (0.6) |
| Anemia in neoplasms | 2 (0.6) |
| Axial spondyloarthritis | 2 (0.6) |
| Conduct disorder | 2 (0.6) |
| Contraception | 2 (0.6) |
| Juvenile idiopathic arthritis | 2 (0.6) |
| Leiomyosarcoma | 2 (0.6) |
| Liposarcoma | 2 (0.6) |
| Psoriatic arthritis | 2 (0.6) |
| Sarcoidosis | 2 (0.6) |
| Acne vulgaris | 1 (0.3) |
| Anemia in acquired immunodeficiency syndrome | 1 (0.3) |
| Anemia in multiple myeloma | 1 (0.3) |
| Anxiety disorder | 1 (0.3) |
| Asthma | 1 (0.3) |
| Atopic dermatitis | 1 (0.3) |
| Atrial fibrillation | 1 (0.3) |
| B-cell chronic lymphocytic leukemia | 1 (0.3) |
| Binge-eating disorder | 1 (0.3) |
| Breast neoplasm | 1 (0.3) |
| Cachexia | 1 (0.3) |
| Chronic lymphocytic leukemia | 1 (0.3) |
| Chronic thromboembolic pulmonary hypertension | 1 (0.3) |
| Diabetes mellitus, adult onset | 1 (0.3) |
| Healthy volunteers | 1 (0.3) |
| Helminth infection | 1 (0.3) |
| Hypertension | 1 (0.3) |
| Mantle cell lymphoma | 1 (0.3) |
| Melanoma | 1 (0.3) |
| Myxoid liposarcoma | 1 (0.3) |
| Niemann-Pick disease, type C | 1 (0.3) |
| Ovarian neoplasm | 1 (0.3) |
| Partial seizure disorder | 1 (0.3) |
| Psychosis | 1 (0.3) |
| Pulmonary fibrosis in systemic sclerosis | 1 (0.3) |
| Raynaud's phenomenon in systemic sclerosis | 1 (0.3) |
| Seizure | 1 (0.3) |
| Small lymphocytic lymphoma | 1 (0.3) |
| Solid tumor | 1 (0.3) |
| Urinary tract infection, pyelonephritis | 1 (0.3) |
| Venous thrombosis | 1 (0.3) |
